# The Role Of Serum Electrolyte Level In Stroke Prognosis

**DOI:** 10.1101/2022.07.24.22277969

**Authors:** Onjal Taywade, Neeta Chourasiya, Monali Hiwarkar, Anup Nillawar

**Affiliations:** Dept of Biochemistry All India Institute of Medical Sciences, Bilaspur, HP, India; Dept of Biochemistry, LNCTMC&SH, Indore, MP, India; Dept Of Anatomy, All India Institutes of Medical Sciences, Bilaspur, HP, India, Mob 8003376962.; Department of Biochemistry, BKL Walawalkar Rural medical College, Sawarde, Chiplun, Maharashtra, India

**Author notes:** **Previous affiliation where work is carried out:** Dept of Biochemistry, Pacific Medical College and Hospital, Udaipur, India (Onjal Taywade, Neeta Chourasiya). **Previous Affiliation:** Department of Anatomy, Pacific Medical College and Hospital, Udaipur, India (Monali Hiwarkar).

**Keywords:** Stroke, NIHSS, MRS-3, Hyponatremia, Serum electrolytes

## Abstract

**Background and introduction:** Acute stroke is one of the common medical emergencies in India that contribute to mortality as well as morbidity. NIHSS scale is still the best clinical conglomerate for initial evaluation of stroke, while Modified Rankin scale performed at 3 months is a simple scale to gauge long-term neurological deficit. Hyponatremia is a common accompanying electrolyte disturbance with short as well as long-term implications in stroke.

**Aim and objectives:** With this background, a study with the following objectives was planned: 1. To estimate the incidence of electrolyte disturbance in acute ischemic stroke, 2. To determine the association of electrolyte disturbances with clinical assessment scales like NIHSS scale, ASPECT score, and MRS-3-month score.

**Material and Methods:** A prospective observational study was conducted on forty-five patients presenting with acute ischemic stroke diagnosed on imaging studies and evaluated for electrolytes (Na^+^, K^+^, Cl^-^).

**Results:** 55% patients had some or other electrolyte disturbance, hyponatremia(33%) being the most common, while hyperchloremia(15%) was the second. The NIHSS score showed better correlation with MRS score than ASPECT score or electrolyte disturbances.

**Conclusion:** Hyponatremia was found to be the most common electrolyte disturbance in acute ischemic stroke. This study attempted to underscore the role of electrolyte disturbance in the prognostication of acute stroke. As electrolyte measurement is feasible in low resource settings; its relevance and utility are even more pronounced.

## Introduction

Cerebrovascular accident or stroke are characterised by the death of brain cells due to lack of oxygen following the interruption in the blood supply to brain parenchyma. The aetiology being ischemia or hemorrhage, either due to the blockage or rupture of arteries supplying blood to the brain tissue. It is the second most common cause of death and the third leading cause of disability worldwide(1). In India, the cumulative incidence of stroke ranges from 105 to 152/100,000 persons per year, and the crude prevalence of stroke ranges from 44.29 to 559/100,000 persons in different parts of the country which is higher than in developed countries(2). There is significant mortality and morbidity associated with stroke with almost half of the survivors being disabled for the rest of life(3).

The severity of stroke was assessed clinically using National Institutes of Health Stroke Scale (NIHSS). It’s 42-point scale that quantifies neurologic deficits in 11 categories. NIHSS score >16 is considered as a marker of poor outcome and NIHSS score of 0-16 indicates, good prognosis (3).

The neurological outcome was measured by the modified Rankin scale (MRS) score at 3 months from the onset of symptoms. MRS is a function assessment scale to assess the neurological deficit, where a score of zero indicates absence of symptoms while a score of 5 indicates severe disability. MRS score 0-2 was considered as a good outcome while 3-6 was considered as poor outcome(4).

ASPECTS score (Alberta Stroke Program Early CT Score) is a 10-point quantitative topographic CT scan score used in patients with middle cerebral artery (MCA) stroke and were adapted for the posterior circulation. An ASPECTS score less than or equal to 7 predicts a worse functional outcome at 3 months(5).

Serum electrolyte disturbances have been observed in both ischemic as well as in hemorrhagic stroke. Hyponatremia is the most common serum electrolyte imbalance seen in acute cerebrovascular insults(6). Disturbance in serum levels of both sodium (hyponatremia or hypernatremia) and potassium could be seen in a patient with acute stroke(7). The utility of serum electrolyte levels as a marker to assess the severity and prognosis of stroke has not yet been fully established. The present study attempts to address this aspect.

## Material and Methods

This was a prospective observational study which was conducted in the department of Biochemistry, over twelve months. (July 2016 to July 2017) after getting approval from Institutional Ethics Committee.

In this study, we enrolled patients who were admitted in the hospital with varying degrees of stroke. As patients were triaged with the help of clinical scales and ASPECT scores on imaging, they underwent intervention according to standard guidelines. We included only ischemic stroke patients and excluded hemorrhagic infarcts. Patients with head injury, brain tumour, gastroenteritis, pulmonary Koch’s, malignancy, history of drugs causing hyponatremia, or recent surgery were excluded. Patients who succumbed within 3 months of stroke were also excluded. Forty-five stroke patients were enrolled within twelve months period whose data was analysed.

We analysed serum electrolytes (Na^+^, K^+^, and Cl^-^) at the time of admission on ‘Roche 9180’ electrolyte analyser. The results were correlated with NIHSS admission score and MRS-3-month score. We tried to throw light on the spectrum of electrolyte disturbances in stroke patients and their likely association with MRS 3-month score.

### Statistical analysis

The data for Na^+^, K^+^, and Cl^-^ levels was compiled in the excel sheet, while risk factor conditions were reported as count(n) and percentage(%) (Table 1). To assess the statistical significance, t-test was employed. Correlation was assessed using Spearman Rank test after testing the data for normality. All tests were carried out at a 5% level of significance. The statistical analysis of the data was done using SPSS version 16 (SPSS Inc., Chicago).

**Table 1:** Demographic and clinical details of the recruited stroke patients:

| **Demographics** | **Ischemic Stroke (on imaging)**<br>**Cases (n=45)** |
| --- | --- |
| Age in years (Interquartile range) | 60.28(20) |
| Gender-Male (%) | 30(66.6%) |
| **Risk factors** | **Number (%)** |
| Hypertension | 30(66.6%) |
| Diabetes mellitus | 20(44.4%) |
| Hyperlipidemia | 30(66.6%) |
| Smoking | 20(44.4%) |
| Alcoholic | 25(55.5%) |

## Results

Our results showed that 25 (55%) patients showed some or other electrolyte disturbance. Amongst this, hyponatremia (Na^+^<135mEq/L) is the commonest being 33% of patients presented with hyponatremia and another 15% patients showed hyperchloremia (Cl^-^ >110mEq/L) and above) and hyperkalemia (K^+^>5 mEq/L) was found in 10% of patients. Thus, in total, 25 out of 45 patients showed some or other electrolyte disturbance, either in single or multiple electrolytes. Hypernatremia and hypokalemia were quite uncommon. (Table 2)

**Table 2:** Showing the proportion of patients with electrolyte disturbances:

| **Sr No** | **Electrolyte Abnormality** | **Proportion of cases (%)** |
| --- | --- | --- |
| 1 | Hyponatremia ($\text{Na}^+ < 135 \text{ mEq/L}$) | 33 |
| 2 | Hypernatremia ($\text{Na}^+ > 145 \text{ mEq/L}$) | 00 |
| 3 | Hypochloremia ($\text{Cl}^- < 95 \text{ mEq/L}$) | 04 |
| 4 | Hyperchloremia ($\text{Cl}^- \geq 110 \text{ mEq/L}$) | 15 |
| 5 | Hypokalemia ($\text{K}^+ < 3.5 \text{ mEq/L}$) | 02 |
| 6 | Hyperkalemia ($\text{K}^+ > 5 \text{ mEq/L}$) | 10 |
| 7 | At least One or More electrolyte disturbance | 55 |

We quantified the outcome of stroke with MRS-3-month score (On 0 to 5 scales, 5 being the worst outcome). When we tried to correlate this score with parameters of presentation like NIHSS and ASPECT score and electrolytes, we got the results represented in graph 1.

This shows that the clinical conglomerate indicator NIHSS score is having the better correlation with MRS-3 score than ASPECT score or electrolyte disturbances. (Table 3) But certainly, electrolyte disturbance poses an additional risk affecting the outcome.

**Table 3:**
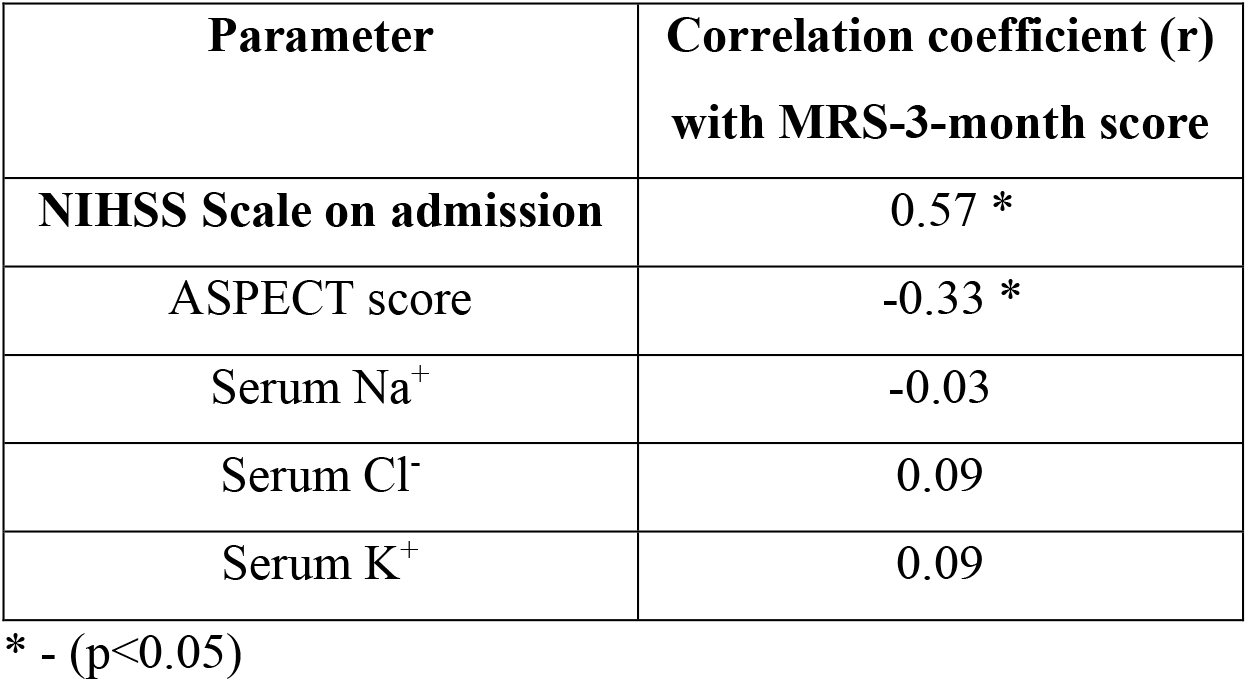
Correlation of MRS-3-months Score (0-5 on scales) with other variables including Electrolytes:

| **Parameter** | **Correlation coefficient (r)**<br>**with MRS-3-month score** |
| --- | --- |
| **NIHSS Scale on admission** | 0.57 * |
| ASPECT score | -0.33 * |
| Serum Na<sup>+</sup> | -0.03 |
| Serum Cl<sup>-</sup> | 0.09 |
| Serum K<sup>+</sup> | 0.09 |

\* - (p<0.05)

On detailed scrutiny of the results, we found that 4 (10%) patients with NIHSS admission scored less than 6 (which is otherwise considered a safe cut-off) and ASPECT score not less than 8 (which is considered a safe cut-off) were showing bad recovery at the end of 3 months and all these patients were having some or other electrolyte disturbance. (Table 4) This clearly underlines the importance to factor electrolyte disturbance while triaging stroke patients. However, this needs to be ascertained with a larger sample size.

**Table 4:** List of patients with poor outcome at 3 months MRS score and presence of electrolyte disturbance:

| **Sr.**<br>**No** | **NIHSS**<br>**Admission**<br>**score** | **Na<sup>+</sup>**<br>**mEq/L** | **K<sup>+</sup>**<br>**mEq/L** | **Cl<sup>-</sup>**<br>**mEq/L** | **MRS-3-**<br>**month**<br>**score** | **ASPECT**<br>**score** |
| --- | --- | --- | --- | --- | --- | --- |
| 1 | 9 | 137 | 4.7 | 113 | 4 | 8 |
| 2 | 4 | 136 | 4.5 | 109 | 4 | 9 |
| 3 | 6 | 119 | 5.2 | 92 | 3 | 8 |
| 4 | 6 | 140 | 4.4 | 112 | 4 | 8 |

## Discussion

Hyponatremia remains the most common electrolyte abnormality seen in hospitalized patients and is associated with increased all-cause mortality, although a cause and effect relation is not always established.(8)(9) Electrolyte imbalance, especially hyponatremia(HN) in acute stroke (commonly defined as serum sodium levels <135mmol/L) of both ischemic and haemorrhagic aetiologies is well documented. The reported incidence varying from 8 to 30% on admission.(9) Metanalysis by Liamis et al, showed varying prevalence of HN in IS on admission from 6.3 to 15.2%.(10) Hyponatremia is more frequently seen in haemorrhagic (HS) than in ischemic stroke (IS). Study by Gala-Błądzińska et al (11) showed prevalence of hyponatremia 22% in HS and 18 % in IS. In the same study, the correlation with NIHSS scale on admission showed a positive correlation with HN and it was pointing towards a bad prognosis. However, our study did not show any correlation between HN and NIHSS score on admission.

Furthermore, we attempted to find a relation between electrolyte imbalance on admission and MRS-3-month score. Though we did not find any statistical association in between these two, we found 4 (10%) peculiar patients (Table 4). Although in the present study NIHSS score correlated better with MRS-3-month score than serum electrolyte levels, four of our patients with deranged MRS-3-month score had serum electrolyte disturbances with normal NIHSS scores. This underlines the importance of serum electrolyte imbalance as a possible prognostic marker in a stroke patient. Previous few studies point towards the prognostic value of estimating serum electrolyte levels in stroke patients(6)(12)(13). Hence, serum electrolyte levels being the baseline investigation should be used to assess the severity and prognosis of stroke.

Hyponatremia is one of the important causes of persistent altered sensorium in stroke patients. There are many precipitating factors for hyponatremia in stroke like dietary restriction of sodium for controlling hypertension, infections and medications viz analgesics, antidepressants, barbiturates, carbamazepine, proton pump inhibitors, antibiotics, oral hypoglycemics etc. The finding of abnormal sodium levels should always prompt specific investigation into the underlying cause. In a few studies, hyponatremia in the acute stroke stage was seen up to 15% of cases and has been found to be a predictor of 3-year mortality in patients with acute first-ever ischemic stroke, while few others found hyponatremia incidence up to 35% approximately(10). The exact mechanism for hyponatremia in stroke is unclear, but one hypothesis states that it is due to an exaggerated renal pressure natriuresis that occurs from increased sympathetic nervous system activity. New onset hyponatremia in acute stroke is most commonly the result of excessive and inappropriate secretion of anti-diuretic hormone (ADH) or cerebral salt wasting syndrome (CSWS). In SIADH, there is excessive release of ADH leading to preferential water retention over sodium leading to hyponatremia. And in CSWS, due to neuronal damage and associated excessive release of atrial natriuretic peptide(ANP), there is large urinary loss of Na(14). Table 5 can be used to differentiate in between SIADH and CSWS(14)(15). Although we applied this table to delineate these two causes of hyponatremia, it is likely that the small sample size did not yield much significant results and outcome. The symptoms directly attributable to hyponatremia primarily occur with acute reductions in the plasma sodium level. Generally, nausea and malaise are the earliest findings and are commonly seen when the plasma sodium concentration falls below 125-130 mEq/L. This may be followed by headache, lethargy, even drowsiness, seizures, coma, and respiratory arrest if the plasma sodium concentration falls below 115-120 mEq/L. Hyponatremia has been found to be an independent predictor of deaths in middle-aged and elderly population even in the absence of stroke, cardiovascular disease, or cancer. (9) Hence, the initial incidental finding of an abnormal sodium level should always prompt specific investigation into the underlying for optimal management.

**Table 5:** Laboratory markers to differentiate between CSWS and SIADH in cases of Hyponatremia:

| **Clinical/ Lab Parameter** | **CSWS** | **SIADH** |
| --- | --- | --- |
| Hematocrit | Increased | Decreased |
| BUN/Creatinine | Increased | Decreased |
| Serum Uric acid | Normal or increased | Decreased |
| Serum Albumin | Increased | Normal |
| Treatment | Normal Saline | Fluid restriction |

In our study, the second most common electrolyte abnormality was hyperchloremia cut off taken as ≥ 110mEq/L. Those patients having good NIHHS score on admission, but later deteriorated at 3 months, MRS invariably had hyperchloremia, but an independent association could not be found. Some of the past studies found similar trends where hyperchloremia was associated with bad long term outcome in stroke(16)(17). Causes of hyperchloremia need to be looked further as any excessive use of intravenous infusions, presence of normal anion gap metabolic acidosis, or acute kidney injury and an attempt should be made to determine the causal relationship between hyperchloremia and poor outcome, if any(18). Further studies are also suggested to elucidate the precise role of serum electrolyte level in stroke. Electrolyte measurement is technically easy and feasible even in resource limiting places, so it has enormous utility. Electrolyte disturbances as such are common in elderly population, especially those with associated comorbidities. Hence, it is advisable to screen stroke patients for electrolyte imbalance at the time of admission and monitor them for complications during the course of time. Moreover, it will be prudent to see whether the appropriate restoration of electrolyte levels improves outcome in patients with acute stroke.

## Conclusion

This study highlights the role of electrolyte disturbances in the prognostication of acute stroke, especially in patients where the clinical scale score might be misleading. Hyponatremia is found to be the most common electrolyte disturbance, followed by hyperchloremia in acute ischemic stroke. Stroke patients with poor MRS-3-month score, showed the presence of electrolyte disturbance at the onset of acute stroke. Thus, base-line electrolyte assessment could have a precise and larger role in the prognostication of acute stroke which needs further evaluation.

## Limitations of the study

As multiple, interdependent variables are involved in affecting serum electrolytes in acute stroke, a larger multicentric cohort study is suggested to assess the association of electrolytes in short as well as long-term outcomes in acute stroke.

## Data Availability

All data produced in the present study are available upon reasonable request to the authors

## Acknowledgement

We thank all our patients and the Department of Neurology, Pacific Medical College and Hospital, Udaipur to support this study.

---

**Graph 1:**
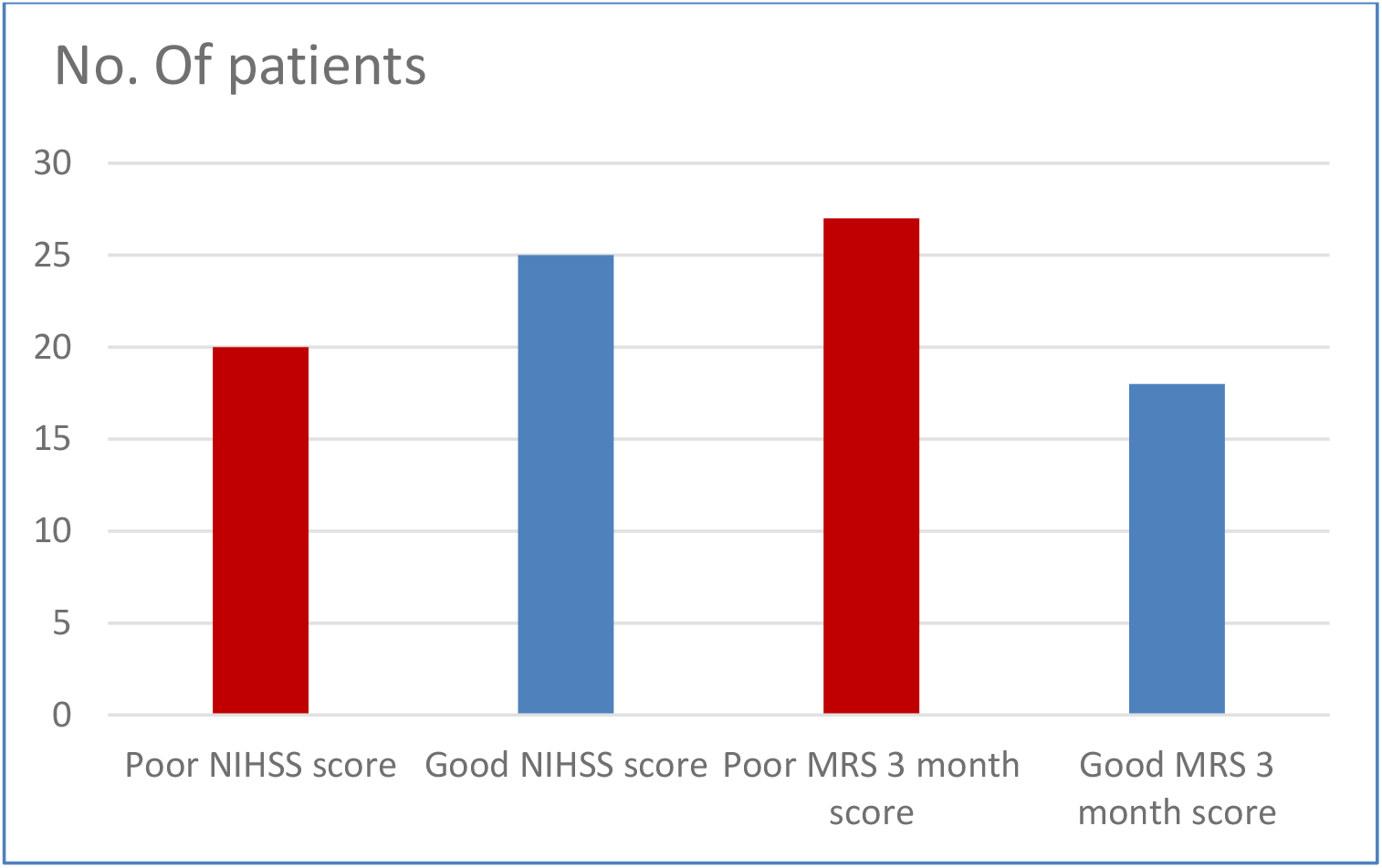
Distribution of patients according to NIHSS scale and MRS-3-month score:

